# A Multi-Ancestry Polygenic Risk Score for Coronary Heart Disease Based on an Ancestrally Diverse Genome-Wide Association Study and Population-Specific Optimization

**DOI:** 10.1101/2023.06.02.23290896

**Authors:** Johanna L. Smith, Catherine Tcheandjieu, Ozan Dikilitas, Kruthika Iyer, Kazuo Miyazawa, Austin Hilliard, Julie Lynch, Jerome I. Rotter, Yii-Der Ida Chen, Wayne Huey-Herng Sheu, Kyong-Mi Chang, Stavroula Kanoni, Phil Tsao, Kaoru Ito, Matthew Kosel, Shoa L. Clarke, Daniel J. Schaid, Themistocles L. Assimes, Iftikhar J. Kullo

## Abstract

**Background:** Predictive performance of polygenic risk scores (PRS) varies across populations. To facilitate equitable clinical use, we developed PRS for coronary heart disease (PRS_CHD_) for 5 genetic ancestry groups.

**Methods:** We derived ancestry-specific and multi-ancestry PRS_CHD_ based on pruning and thresholding (PRS_P+T_) and continuous shrinkage priors (PRS_CSx_) applied on summary statistics from the largest multi-ancestry genome-wide meta-analysis for CHD to date, including 1.1 million participants from 5 continental populations. Following training and optimization of PRS_CHD_ in the Million Veteran Program, we evaluated predictive performance of the best performing PRS_CHD_ in 176,988 individuals across 9 cohorts of diverse genetic ancestry.

**Results:** Multi-ancestry PRS_P+T_ outperformed ancestry specific PRS_P+T_ across a range of tuning values. In training stage, for all ancestry groups, PRS_CSx_ performed beter than PRS_P+T_ and multi-ancestry PRS outperformed ancestry-specific PRS. In independent validation cohorts, the selected multi-ancestry PRS_P+T_ demonstrated the strongest association with CHD in individuals of South Asian (SAS) and European (EUR) ancestry (OR per 1SD[95% CI]; 2.75[2.41-3.14], 1.65[1.59-1.72]), followed by East Asian (EAS) (1.56[1.50-1.61]), Hispanic/Latino (HIS) (1.38[1.24-1.54]), and weakest in African (AFR) ancestry (1.16[1.11-1.21]). The selected multi-ancestry PRS_CSx_ showed stronger association with CHD in comparison within each ancestry group where the association was strongest in SAS (2.67[2.38-3.00]) and EUR (1.65[1.59-1.71]), progressively decreasing in EAS (1.59[1.54-1.64]), HIS (1.51[1.35-1.69]), and lowest in AFR (1.20[1.15-1.26]).

**Conclusions:** Utilizing diverse summary statistics from a large multi-ancestry genome-wide meta-analysis led to improved performance of PRS_CHD_ in most ancestry groups compared to single-ancestry methods. Improvement of predictive performance was limited, specifically in AFR and HIS, despite use of one of the largest and most diverse set of training and validation cohorts to date. This highlights the need for larger GWAS datasets of AFR and HIS individuals to enhance performance of PRS_CHD_.

## Introduction

Coronary heart disease (CHD) is a leading cause of death in the United States (U.S.) and worldwide ^1^. CHD has an estimated heritability of 40-60% and the majority of the heritable risk is atributable to a polygenic component, i.e., the aggregation of modest effects across many genetic variants ^2^. Polygenic risk scores (PRS) capture a proportion of that heritability and are typically constructed by summing the products of the effect-size and the number of risk alleles at associated loci ^3,4^. PRS for CHD have evolved over the last decade as progressively larger genome wide association studies (GWAS) have been reported ^5-8^. These PRS have been evaluated in several studies and are associated with incident CHD independent of conventional risk factors such as hypertension, hypercholesterolemia, diabetes, and smoking as well as family history of CHD ^8-10^.

Most PRS for CHD have been developed, optimized, and validated in cohorts consisting largelyof individuals of European (EUR) ancestry (here and throughout the manuscript ‘ancestry’ refers to genetic ancestry) ^11-14^. Furthermore, the portability of these PRS to non-EUR groups is impacted by differences in allele frequencies (AF), effect sizes, and linkage disequilibrium (LD) paterns across ancestry groups, typically resulting in reduced predictive performance as studied populations diverge in these factors; an observation most notable between EUR and African (AFR) ancestry populations ^6,11,15^. We previously observed significantly lower performance of several EUR-derived PRS for CHD in AFR ancestry individuals ^16,17^. To prevent exacerbation of health disparities in the context of genomic medicine, there is a need to improve performance of PRS for CHD for non-EUR populations.

In this study, we leveraged a large scale, ancestrally diverse genome-wide meta-analysis for CHD to construct PRS for CHD optimized for EUR, AFR, Hispanic/Latino (HIS), East Asian (EAS), and South Asian (SAS) ancestries. To this end, we utilized two PRS derivation methods, pruning and thresholding (P+T) and the continuous shrinkage prior based PRS-CSx ^8,18^. We assessed the performance of the multi-ancestry PRS in individuals with diverse ancestry belonging to 8 independent validation cohorts. Finally, a PRS was selected for clinical implementation in the electronic Medical Records and Genomics (eMERGE) network phase IV study in which PRS-informed risk profiles for several common conditions are being returned to participants ^19^.

## Methods

### GWAS Summary Statistics for PRS Development

We developed PRS using both ancestry-specific and multi-ancestry meta-analysis summary statistics from a large-scale multi-ancestry GWAS for CHD including 1.1 million diverse participants with 243,392 CHD cases ^17^. This diverse meta-analysis included 17,202 AFR, 6,378 HIS, 29,319 EAS, and 190,776 EUR individuals with CHD belonging to four cohorts including the Million Veteran Program (MVP), the UK Biobank (UKBB), CARDIoGRAMplusC4D Consortium (2015 release), and Biobank Japan (BBJ) (Figure 1) ^17,20-22^.

**Figure 1.**
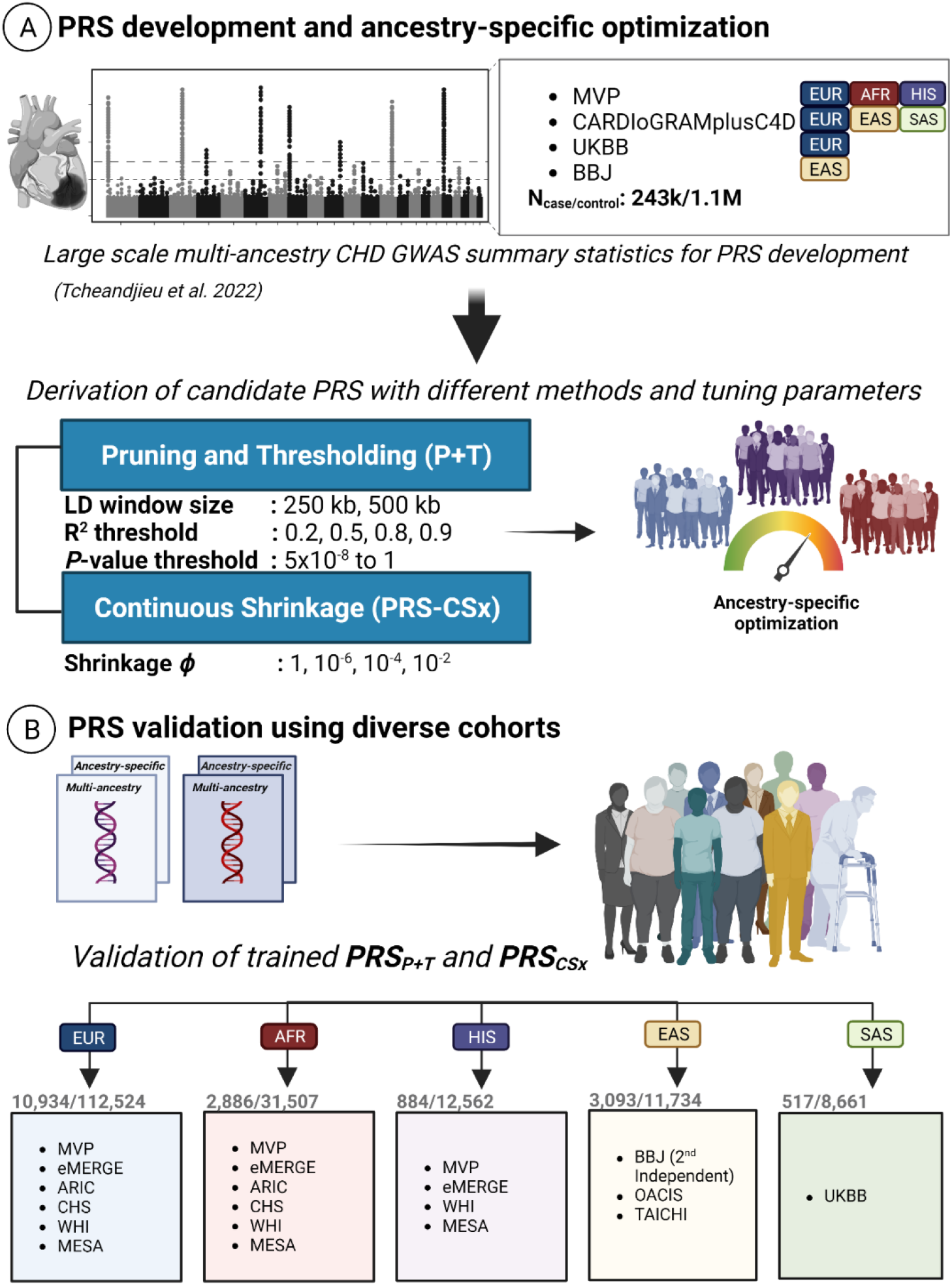
Polygenic Risk Score development using independent MVP cohorts of diverse ancestry.

We used two distinct methods to construct PRS, namely, pruning and thresholding (P+T) and the continuous shrinkage prior based PRS-CSx ^8,18^. Ancestry-specific PRS were defined from ancestry-specific GWAS summary statistics (i.e., EUR specific summary statistics were used to develop a EUR specific PRS), and multi-ancestry PRS were defined as PRS derived from multi-ancestry summary statistics. These PRS were then trained and optimized in a separate set of individuals from the MVP and externally validated in several diverse cohorts including the Atherosclerosis Risk in Communities (ARIC) ^23^, Multi-Ethnic Study of Atherosclerosis (MESA) ^24^, Cardiovascular Health Study (CHS) ^25^, Women’s Health Initiative (WHI) ^26^, eMERGE Phases I-III genotyped cohort ^27^, Biobank Japan (BBJ) ^28^, Osaka Acute Coronary Insufficiency (OACIS) study ^29^, the TAICHI Consortium ^30^, and individuals of SAS ancestry from the UKBB ^31^ (Table S1; Supplemental File 1).

### Pruning and Thresholding (P+T)

We derived two independent sets of PRS (ancestry-specific and multi-ancestry PRS) in two sequential steps: First, we excluded from the base GWAS summary statistics, correlated single nucleotide variants (SNVs) by LD pruning, applying 4 different *R*^2^ thresholding values (0.2, 0.5, 0.8, and 0.9) and 2 different window distances (250kb and 500kb) within which these *R*^2^ were applied. LD pruning for ancestry-specific PRS was performed based on reference panels comprised of 4,000 participants from each respective ancestry (EUR, AFR, HIS, and ASN), selected among MVP participants included in the large-scale GWAS for CHD. The LD pruning for the multi-ancestry PRS was performed on the full subset of 16,000 individuals from EUR, AFR, HIS, and ASN as the reference panel. This step generated 8 ancestry-specific summary statistics and 8 multi-ancestry summary statistics for PRS development. Second, for each newly generated summary statistic from step 1, we applied 16 different *p*-value thresholds (5×10^−08^, 1×10^−04^, 0.001, 0.005, 0.01, 0.05, 0.1, 0.2, 0.3, 0.4, 0.5, 0.6, 0.7, 0.8, 0.9, and 1) (Figure S1; Supplemental File 1). These led to 128 summary statistics within each ancestry, which were used to train the ancestry-specific PRS. Similarly, we obtained 128 multi-ancestry-based summary statistics to train the multi-ancestry PRS (PRS_P+T_).

### Continuous shrinkage (PRS-CSx)

We applied a continuous shrinkage method, PRS-CSx (PRS_CSx_), on the effect sizes of a subset of 1.4 million well curated HapMap SNVs on each ancestry-specific summary statistic. To identify the optimal shrinkage parameter, we applied 4 different global shrinkage phi parameters (1, 1*e*^−02^, 1*e*^−04^, and 1*e*^−06^). LD reference panels used were EUR, AFR, AMR and EAS from the 1000 Genomes project. The multi-ancestry PRS were constructed from the meta-analysis of ancestry-specific summary statistics obtained after applying the global shrinkage phi. For each ancestry, 4 ancestry-specific newly derived summary statistics were obtained to train ancestry-specific PRS and 4 newly derived multi-ancestry summary statistics were obtained for train the multi-ancestry PRS (Figure S2; Supplemental File 1). A total of 12 ancestry-specific PRS (one for each global shrinkage parameter value used for each ancestry group and 4 multi-ancestry PRS) were chosen for further development (Figure S3; Supplemental File 1).

### PRS Training

Following the construction of the ancestry-specific and multi-ancestry PRS_P+T_ and PRS_CSx_ across a range of training specifications, we proceeded to assess their performance in an independent set of prevalent cases and controls from the MVP (Figure 1B, PRS Training) using multivariable logistic regression with adjustment for age at CHD event for cases and age at the last visit in the electronic health record (EHR) for controls, year of birth, sex, and the first 5 principal components (PCs). We compared parameter training on the multi-ancestry reference panel set versus population-specific reference panel. Ancestry-specific PRS were evaluated in the corresponding ancestry, whereas the multi-ancestry PRS were evaluated in each ancestry. PRS with the highest observed odds ratio (OR) for CHD per 1 standard deviation (SD) increase were deemed to have the optimal training parameter values across ancestry populations and subsequently advanced for validation.

### PRS Validation in the Million Veteran Program and Additional External Cohorts

Ancestry-specific and multi-ancestry PRS_P+T_ and PRS_CSx_ trained for each genetic ancestry group were validated in an independent cohort from the MVP and several additional diverse cohorts (Figure 1C, Diverse Cohorts for PRS Validation). The MVP validation cohort was restricted to incident cases of CHD occurring after enrollment, and random controls, in a ratio of 1:10 (Figure 1C) as previously described ^17^. Four prospective cohorts, namely ARIC, MESA, CHS, WHI, a subset of the UKBB comprised of individuals of SAS ancestry, and additionally eMERGE Phases I-III, contributed CHD incident cases and controls of EUR, AFR, HIS, and SAS ancestry for PRS validation. Validation for EAS ancestry included individuals from multiple case-control studies, namely Han Chinese participants from Taiwan as a part of the TAICHI consortium, as well as Japanese participants from the BBJ and OACIS studies who were not part of the multi-ancestry discovery GWAS ^30^.

Within MVP, we used diagnosis and procedure codes to identify individuals with any clinical manifestation of CHD as previously described (Supplemental File 1) ^17^. This definition included both ‘hard’ (e.g., myocardial infarction, revascularization) and ‘sotf’ outcomes (e.g., angina, non-invasive study positive for ischemia). In the 4 external validation NHLBI cohorts and the eMERGE cohort, cases were restricted to myocardial infarction and revascularization. Prevalent cases were defined as all other cases meeting diagnosis/procedure code criteria at the time of enrollment. Additional study details are included in Supplemental File 1.

We calculated OR per 1-SD increase in PRS using multivariable logistic regression across all validation cohorts. The dbGaP, eMERGE, and UKBB cohorts were adjusted for genetic ancestry using a continuous correction further defined in the Supplemental File 1 (Figure S4). The two EAS case-control studies were meta-analyzed using a fixed effect inverse-variance weighted model ^32^. For all external validation cohorts, we additionally estimated OR for CHD for participants in the top 5% of PRS distribution compared to the rest, as well as area under the curve (AUC) discrimination statistic.

Calibration was also assessed using the calibration function in the rms package in R to assess portability to cohorts that were not available for meta-analysis (i.e., the non-EAS cohorts) (Figure S5, Supplemental File 2) ^33,34^.

## Results

### PRS Training

#### Pruning and Thresholding (P+T)

Performance of the ancestry-specific and multi-ancestry PRS_P+T_ in each population is shown in Figure 2. The multi-ancestry PRS_P+T_ systematically outperformed ancestry-specific PRS_P+T_ with noticeably higher OR per SD except for the HIS ancestry group where the performance was similar (Figure 2, Supplemental Figure S2). The multi-ancestry PRS_P+T_, performed best in HIS population, followed by the ASN population (1.78 and 1.73 OR per SD, respectively) (Supplemental File 2). Prediction performance of the PRS_P+T_ for each ancestry was optimal at different *p*-value thresholds (Figure 2, Supplemental Figure S2). The multi-ancestry PRS_P+T_ performed best at *R*^2^ ≤ 0.8 with LD blocks of 250 kb, *p*-value threshold of 0.01 for AFR, 0.03 for EUR, and 0.30 for HIS. However, the differences between these PRS and the PRS optimized at *R*^2^ ≤ 0.8 and a *p*-value = 0.01 were marginal, and the multi-ancestry PRS with a *p*-value threshold of 0.01 was chosen for validation in additional external cohorts.

**Figure 2.**
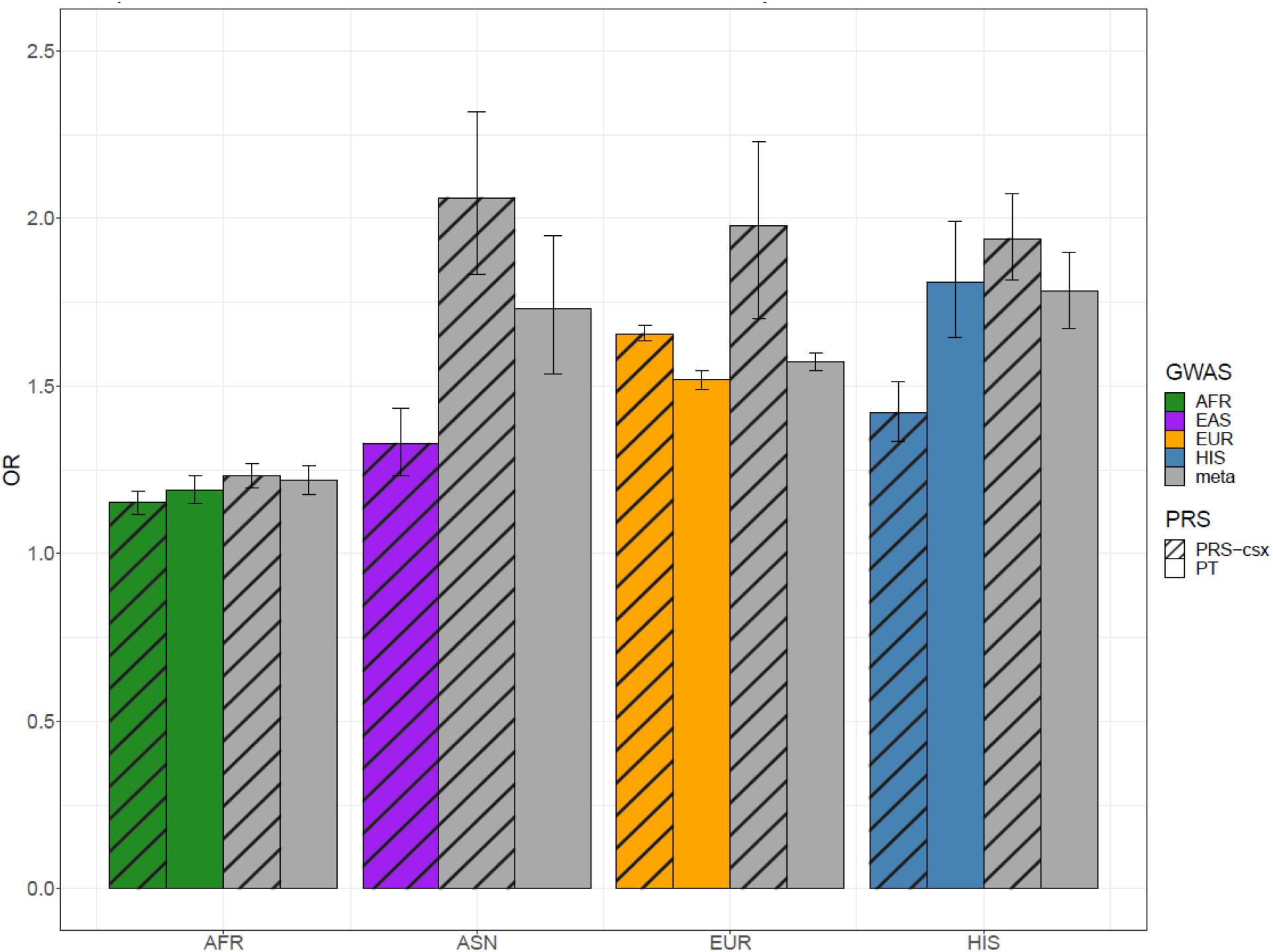
Performance of PRS-CSx (solid bars) or P+T (dashed bars) across genetic ancestry groups when utilizing the diverse MVP training cohort. The colors represent the GWAS summary statistics used to construct the PRS (green for AFR, purple for EAS, orange for EUR, and grey for the multi-ancestry meta-analysis). The Odds Ratios (ORs) per 1 standard deviation (SD) increase with confidence intervals (CIs) in the PRS are represented on the Y-axis and the populations on which the PRS is trained are on the X-axis.

### Continuous shrinkage (PRS-CSx)

The performances of the 12 ancestry-specific PRS_CSx_ and 3 multi-ancestry PRS_CSx_ built using EUR, AFR, HIS, and EAS summary statistics at various global shrinkage phi values for tuning (1*e*^−02^, 1*e*^−04^, and 1*e*^−06^) are shown in Figure 2. For all ancestry groups, *phi* = 1*e*^−02^ resulted in the best predictive performance for PRS_CSx_ and the multi-ancestry PRS outperformed ancestry-specific PRS at this phi value. For the EUR population, both the EUR-derived PRS and the multi-ancestry PRS performed similarly, but ASN and HIS populations performed best with the EUR-derived PRS, while the AFR population performed best with the multi-ancestry PRS (Figure 2, Supplemental File 2). Overall, the multi-ancestry PRS_CSx_ for the ASN population resulted in the highest OR per/SD increase followed by EUR and HIS populations where the strength of association was similar, and lowest in the AFR ancestry.

### PRS Validation

#### Million Veteran Program

Ancestry specific PRS_P+T_ predictive performance (OR per 1 -SD increase) for EUR (1.52), AFR (1.19), and HIS (1.81) was compared to the ancestry-specific PRS_CSx_ performance for EUR (1.66), AFR (1.15), HIS (1.42), and ASN (1.32) (Figure 2; Supplemental File 2). This was also compared to the multi-ancestry-based methods using the same PRS training, i.e., the multi-ancestry PRS_P+T_ for EUR (1.57), AFR (1.22), HIS (1.78), and ASN (1.73), as well as PRS_CSx_ for EUR (1.98), AFR (1.23), HIS (1.94), and ASN (2.06) (Figure 2; Supplemental File 2). Of all the methods assessed at this step, the best performing methods tended to be the multi-ancestry PRS_CSx_ and multi-ancestry PRS_P+T_. However, there were overlapping confidence intervals (CIs) with some single ancestry methods and the single-ancestry PRS_CSx_ for EUR performed well in other ancestries, so we decided to further assess the three methods (Figure 2).

We advanced the ancestry optimized PRS_P+T_ and PRS_CSx_, for validation in an independent set of incident cases and matching controls in ancestry groups of EUR, AFR, HIS, EAS, and SAS individuals.

Predictive performances of the multi-ancestry PRS were assessed within each ancestry group in reference to a previously reported genome-wide PRS (i.e., PRS_metaGRS_ ^10^) constructed using a cohort of predominantly of EUR ancestry (Figure 3) ^17^. In this independent validation cohort, the multi-ancestry PRS_P+T_ and PRS_CSx_ had a higher predictive performance compared to metaGRS (Figure 3). The multi-ancestry PRS_CSx_ had a relative increase in the estimated OR per 1-SD of 12% and 23% in reference to PRS_P+T_ and PRS_metaGRS_, respectively, averaged across all three genetic ancestries.

**Figure 3.**
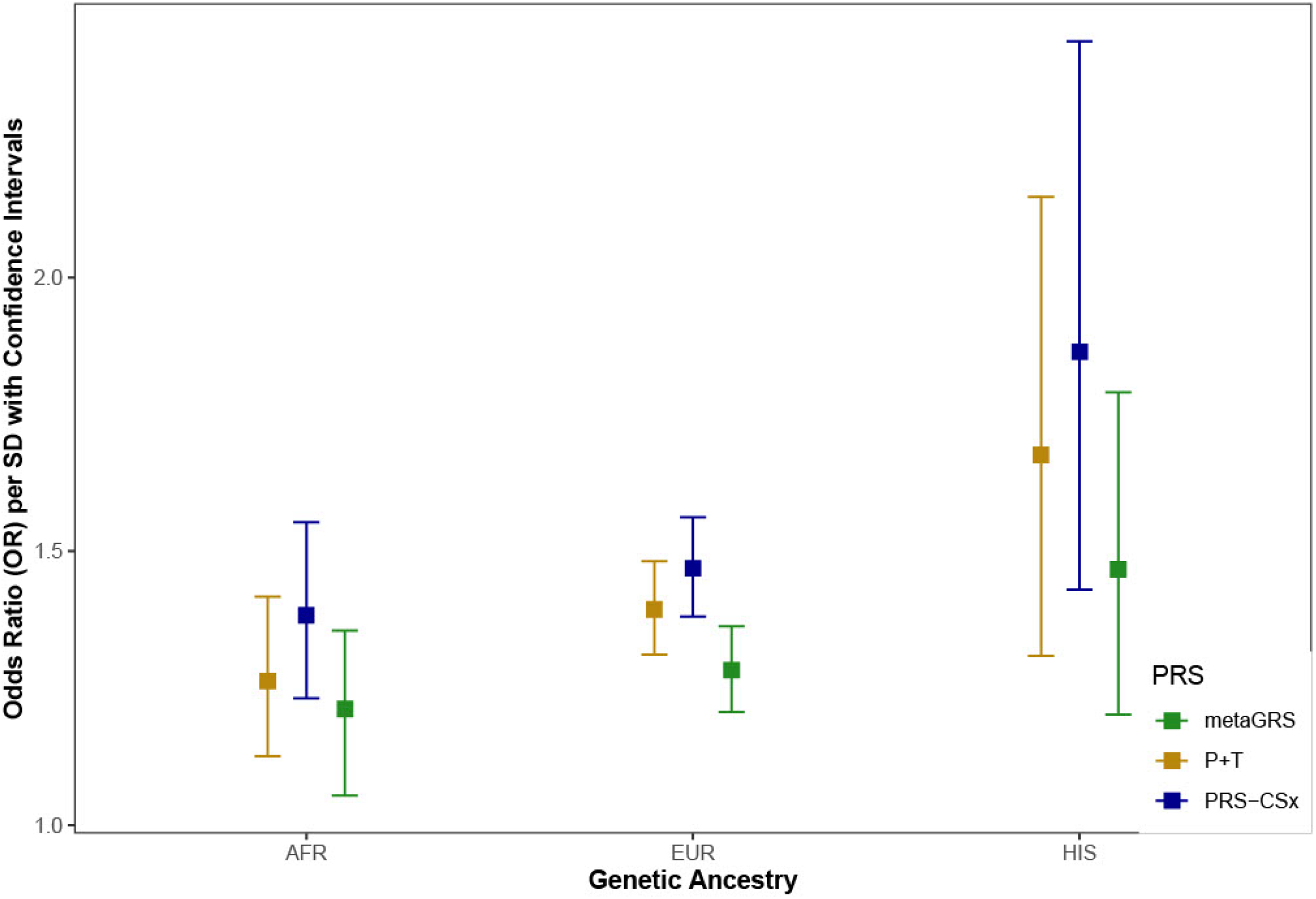
Comparison of a prior PRS (metaGRS) and two new PRS using multi-ancestry summary statistics for the prediction of coronary heart disease (CHD) using the ancestrally diverse training cohort of the MVP. Odds Ratios (ORs) per standard deviation (SD) with confidence intervals (CIs) are shown for each genetic ancestry group as determined in the methods as a result of metaGRS, P+T, and PRS-CSx PRS methods being performed on the MVP training cohort.

### Additional External Validation Cohorts

The best performing PRS_P+T_ were further validated in several additional cohort and case-control studies of CHD including EUR, AFR, HIS, EAS, and SAS participants (Table 1). ORs for ancestry-specific and multi-ancestry PRS_P+T_ ranged from 1.16 in AFR to 2.75 in SAS and were comparable to published reports, despite inclusion of the diverse meta-analysis of GWAS (Supplemental File 2) ^6,17,35,36^. All populations had OR estimates for the top 5% vs the rest of the population ≥ 2.16 for PRS_P+T_ except for AFR (1.68).

**Table 1.** Odds Ratios for incident CHD for multi-ancestry PRS_P+T_ and PRS_CSx_ in diverse ancestry cohorts.

| Ancestry | Age <sup>a</sup><br>(mean±SD<sup>b</sup>) | Cases/Controls | Method | AUC<sup>c</sup> | OR<sup>d</sup> (95% CI<sup>e</sup>)<br>per 1 SD | P-value | OR (95% CI) Top<br>5% vs Rest | P-value |
| --- | --- | --- | --- | --- | --- | --- | --- | --- |
| EUR | 52.4 ± 15.5 | 4,970/47,732 | $PRS_{P+T}$ | 0.773 | 1.65<br>(1.59-1.72) | 3.00E-159 | 2.30<br>(2.07-2.56) | 5.71E-55 |
| | | | $PRS_{CSx}$ | 0.774 | 1.65<br>(1.59-1.71) | 5.19E-171 | 2.48<br>(2.23-2.77) | 5.88E-61 |
| AFR | 53.4 ± 14.7 | 1,359/15,649 | $PRS_{P+T}$ | 0.735 | 1.16<br>(1.11-1.21) | 2.86E-12 | 1.68<br>(1.39-2.03) | 5.51E-08 |
| | | | $PRS_{CSx}$ | 0.736 | 1.20<br>(1.15-1.26) | 7.46E-14 | 1.74<br>(1.41-2.15) | 3.03E-07 |
| HIS | 54.8 ± 14.3 | 314/5,824 | $PRS_{P+T}$ | 0.699 | 1.38<br>(1.24-1.54) | 5.89E-09 | 2.16<br>(1.47-3.19) | 1.02E-04 |
| | | | $PRS_{CSx}$ | 0.706 | 1.51<br>(1.35-1.69) | 7.48E-13 | 2.57<br>(1.77-3.73) | 7.98E-07 |
| EAS | 65.4 ± 12.9 | 6,321/16,430 | $PRS_{P+T}$ | 0.748 | 1.56<br>(1.50-1.61) | 2.97E-146 | 2.47<br>(2.10-2.90) | 1.24E-39 |
| | | | $PRS_{CSx}$ | 0.762 | 1.59<br>(1.54-1.64) | 2.41E-160 | 2.34<br>(2.06-2.66) | 1.78E-28 |
| SAS | 53.2 ± 8.4 | 517/8,661 | $PRS_{P+T}$ | 0.786 | 2.75<br>(2.41-3.14) | 9.44E-52 | 3.95<br>(3.03-5.15) | 3.07E-24 |
| | | | $PRS_{CSx}$ | 0.803 | 2.67<br>(2.38-3.00) | 1.48E-63 | 4.92<br>(3.81-6.35) | 3.90E-34 |

<sup>a</sup> Age- Age at enrollment
<sup>b</sup> SD- Standard Deviation
<sup>c</sup> AUC-Area under the Curve
<sup>d</sup> OR- Odds Ratio
<sup>e</sup> CI- Confidence Interval

The two best performing PRS_CSx_ in the training dataset, a EUR-tuned PRS and a multi-ancestry PRS, both with a tuning global phi value of 1*e*^−02^, demonstrated similar performances in our validation cohorts (Table 1, Table S2; Supplemental File 1) as the multi-ancestry PRS marginally outperformed the EUR-tuned PRS in all but the AFR and HIS cohorts. Point estimates of the OR for subjects in the top 5^th^ percentile of scores compared to the remaining participants shitied trend compared to those observed for the ORs per 1-SD for AFR, HIS, and SAS populations, but these differences were in the context of mostly overlapping 95% confidence intervals. When comparing the multi-ancestry PRS_P+T_ to PRS_CSx_, the point estimates of ORs were similar but higher for the multi-ancestry PRS_CSx_ for EUR, AFR, HIS, and EAS populations. The OR per 1-SD was lower for the multi-ancestry PRS_CSx_ for the SAS population (Table 1).

## Discussion

Using summary statistics from the largest multi-ancestry GWAS meta-analysis for CHD to date and 9 independent validations cohorts, cumulatively comprised of 1.1 million diverse participants including nearly a quarter of a million CHD cases of EUR, AFR, HIS, EAS, and SAS descent ^17^, we developed, trained, and validated multi-ancestry and ancestry-specific PRS models to address the gap in predictive performance that currently exists between EUR and non-EUR ancestries.

We observed that the use of summary statistics from a multi-ancestry GWAS meta-analysis, in comparison to the use of ancestry-specific summary statistics, improved PRS performance in majority of the ancestry groups. PRS that leveraged shared information between ancestries to estimate SNV weights (i.e., PRS_CSx_) modestly outperformed the P+T method (i.e., PRS_P+T_). Based on the multi-ancestry informed PRS_CSx_, individuals in the high-genetic risk group (i.e., top 5% of the PRS distribution) compared to the remaining participants in the respective ancestry group (EUR, AFR, HIS, EAS, and SAS), had 2.5-fold, 1.7-fold, 2.5-fold, 2.3-fold, and 5-fold increased risk of CHD, respectively. These results collectively highlight complementary effects of integrating summary statistics from multiple ancestries and the use of PRS derivation methods that leverage shared information and LD diversity between ancestry groups to improve polygenic risk prediction for CHD.

Although remarkable progress has been achieved to date in both genomic discovery and polygenic risk prediction among EUR cohorts ^5,7-10,37-39^, similar progress has not occurred among non-EUR populations due to their underrepresentation in genomic studies ^11-14^. In recent years, the number of large-scale multi-ancestry GWAS and polygenic risk prediction studies have increased with the establishment of ancestrally diverse biobanks and collaborations efforts ^17,18,30,40-43^. Several multi-ancestry genomic studies, including for glycemic, hematologic and lipid traits as well as disease phenotypes such as type 2 diabetes and CHD, have increased the number of discovered loci, and improved fine-mapping and cross-population polygenic risk prediction with inclusion of non-EUR participants ^17,40-42,44^. Our findings are consistent with these results in that integration of summary statistics from several distinct ancestry groups improved predictive performance of PRS for all ancestries, including EUR descent. One possible explanation for these observations is identification of potential causal variants that are more likely to be shared between ancestries but are obscured by population-specific LD paterns ^14,45^. Another likely contributing factor to improved PRS performance is reduced noise in SNV effect size estimates resulting from both weighted average of population-level estimates and increased total sample size ^46,47^.

Despite the use of the largest ancestrally diverse cohort available to date, the improvement in the predictive performance of PRS_CHD_ was limited in individuals of AFR ancestry compared to other ancestry groups. Prior reports investigating portability of PRS between populations noted that prediction performance across a range of traits and phenotypes ^6,11,15,16,48,49^ decayed with increasing genetic distance between study cohorts. Among the continental ancestry groups included in this study, AFR is the most genetically distant population from EUR and hence the modest increase in prediction performance with a multi-ancestry PRS_CHD_ compared to the ancestry-specific counterpart. A recent report showed similar heritability for CHD in the major continental ancestry groups but absence of two common haplotypes at the 9p21 locus in AFR individuals, which corresponds to the largest effect locus in EUR ancestry individuals ^17^. These findings suggest potentially a larger role of ancestry-specific causal variants in individuals of African origin with regards to heritability for CHD.

Although the strength of association of PRS with CHD varied between ancestry groups, it is important to consider epidemiological differences in CHD risk across these populations. In clinical practice, primary prevention guidelines for CHD use absolute risk estimates for clinical decision making, such as 10-year or lifetime risk of a CHD event ^50^. Individuals are typically classified into different risk groups (e.g., low, borderline, intermediate, high risk) with a correlating intensity of pursued preventive measures. In the United States, African American and South Asian populations have substantially higher atherosclerotic cardiovascular disease (ASCVD) related mortality rates compared to non-Hispanic whites ^1,51^. Therefore, in a future risk model for ASCVD similar to the pooled cohort equation ^52^, incorporation of a PRS for CHD with a narrower risk gradient in African Americans, compared to a much wider gradient in non-Hispanic whites, could have more impact on re-classification into a higher risk group as we have previously shown ^6^.

Implementation of PRS in the clinical seting has begun for CHD, including at Mayo Clinic, where a PRS for CHD is available in the clinical seting, based on the results of the MIGENES clinical trial ^53^. The eMERGE Network, in its phase IV study is returning risk assessments to participants for 11 common conditions, including CHD ^19^. The multi-ancestry PRS_P+T_ for CHD validated in this study ^19^ will be returned to eMERGE participants. One of the major challenges in the clinical use of PRS include variable performance between genetic ancestry populations ^11,15^. Developing robust PRS for diverse ancestry groups is crucial to avoid worsening existing health disparities ^11^ and a National Institute of Health (NIH) funded initiative is addressing this as a priority ^54^. The active recruitment and inclusion of diverse participants and continued development of novel PRS methods that target improvement of cross-population prediction using a variety of approaches (e.g., incorporation of local ancestry ^55^, weighting by trans-ancestry genetic correlation ^56^, and informing by fine-mapping and functional annotation ^57,58^) will be necessary for equitable implementation of PRS. Consequently, we anticipate that PRS for CHD will continue to evolve and improve over time.

### Study Limitations

Despite the large and diverse composition of our study, the external validation for the SAS ancestry was limited to a single cohort with a modest number of cases, reducing the precision of the associated risk estimates. We were not able to include smoking status or family history in the models as the data was not available for all cohorts, and this may have affected the strength of the association of PRS with CHD in our analyses.

## Conclusions

We demonstrated that incorporation of summary statistics from diverse genetic ancestry groups, as opposed to individual ancestry groups alone, and leveraging shared information between these populations, led to improved performance of PRS_CHD_ in majority of the ancestry groups. Despite utilization of one of the largest and most ancestrally diverse set of training and validation cohorts to date, the gain in predictive performance for AFR was limited. Ongoing work is needed to narrow the persistent performance gap for AFR ancestry individuals. Increasing AFR representation at each stage of PRS development is necessary to lessen performance disparities, and such efforts should be a priority for the community of genomics researchers.

## Data Availability

All data produced in the present work are contained in the manuscript

## Acknowledgements

We acknowledge the investigators and participants of the electronic Medical Records and Genomics (eMERGE) Network. Infrastructure for the CHARGE Consortium is supported in part by the National Heart, Lung, and Blood Institute (NHLBI) grant R01HL105756. This work was also supported in part by the National Institutes of Health, National Heart, Lung, Long and Blood Institute (NHLBI) contract 1R01HL151855, R01HL146860, and the National Institute of Diabetes and Digestive and Kidney Diseases contract UM1DK078616.

## Disclosures

### Human and Animal Rights and Informed Consent

This article used data from previously published human studies.

