## Supplemental File 1 for "A Multi-Ancestry Polygenic Risk Score for Coronary Heart Disease Based on an Ancestrally Diverse Genome-Wide Association Study and Population-Specific Optimization"

#### Table of Contents


1. Description of Cohorts and Quality Control (QC)
2. Statistical Adjustments
3. Supplemental Table S1-S2
4. Supplemental Figures S1-S3
5. References


#### Description of Cohorts and Sample Quality Control

The Million Veteran Program (MVP)

The MVP is a U.S. Veteran Affair (VA) national research program, to learn how genes, lifestyle, and military exposures affect health and illness. The MVP was initiated in 2009 and the enrollment started in 2011 with an objective of enrolling over 1 million veterans. Over 900,000 veterans have been enrolled to date from more than 75 VA Medical Centers nationwide. As previously described, informed consent is obtained from all participants to provide blood for genomic analysis and access to their full EHR within the VA prior to and after enrollment including inpatient International Classification of Diseases (ICD9/10) diagnosis codes, Current Procedural Terminology (CPT) codes, clinical laboratory measurements, and reports of diagnostic imaging modalities [1]. We genotyped 650,000 multi-ethnic individuals using a custom Affymetrix Axiom array between 2011 and 2019 in multiple batches. Populations were defined by genetic ancestry using Harmonizing Genetic Ancestry and Self-identified Race/Ethnicity (HARE) [2] and assigned >98% of participants to 1 of 4 non-overlapping groups (EUR, AFR, HIS, and non-HIS ASN). Sex ambiguous individuals were excluded. King v2.0 was used to identify and remove individuals that were at a third-degree relation or closer [3].

To define CAD cases, we used inpatient and outpatient ICD diagnostic and CPT procedure codes to identify participants with clinical CAD in the MVP. EHR data were available retrospectively before enrollment back to October 1999 and prospectively after enrollment until Dec 2020. An individual was classified as a case if he or she had any admission to a VA hospital with a discharge diagnosis of acute myocardial infarction, or any procedure code for revascularization of the coronary arteries, or two or more ICD codes for CAD (410–414) in at least two different encounters. Individuals with only one ICD code for CAD in a single encounter and no discharge diagnoses for acute myocardial infarction or revascularization procedures were excluded from the analyses. The remaining participants were classified as controls.

Prior to imputation, genotyped data was filtered and underwent quality control following Hunter-Zinck et al. 2020. This included removal of genotype missingness > 5% or variants that deviated from expected observed allele frequencies observed in the 1000 Genomes reference sample. After QC, imputation was performed using the 1000 Genomes phase 3 version 5 reference panel Genomes Project Consortium [4] . The full 650,000 genotyped individuals were imputed using EAGLE v2.3 [5] and Minimac3 [6] . Further QC measures were taken as specified in previous publications [1]. After QC, 32 million variants remained among which 12 million SNPs were matched to the summary statistic used to construct the polygenic risk scores [1].

dbGaP Datasets

A brief description of the cohorts chosen from dbGaP for validation is provided below. Complete information regarding the studies are available at https://www.ncbi.nlm.nih.gov/gap/.

The Atherosclerosis Risk in Communities (ARIC) Study [7]

The ARIC study included individuals aged 45-65 years and enrolled from four communities in 1987 with follow up data available through 2016. Follow up was started from the initial examination (visit 1), prevalent cases at baseline were excluded, and incident CHD was defined as a composite of myocardial infarction (MI), fatal coronary event, silent infarction as determined by ECG readings, and having undergone coronary revascularization by the end of available study follow up. dbGaP accession phs0000280.v6.p1.

The Multi-Ethnic Study of Atherosclerosis (MESA) Study [8]

The MESA study included individuals aged 45-84 years from a diverse, population-based sample from six field centers across the US. Follow up was available from the initial visit conducted between 2000-2002 through the end of 2011. Incident CHD events were defined as MI, definite and probable angina if coronary revascularization was performed at the same time or afterwards, resuscitated cardiac arrest, or fatal CHD. DbGap accession phs000209.v2.p1.

The Cardiovascular Health Study (CHS) [9]

The CHS study is a prospective study including individuals aged 65 years and older with initial enrollment in 1989-1990 and a supplemental cohort of predominantly African Americans enrolled in 1992-1993. Study follow up started from the initial visit for all participants with events data available through 2011. Incident CHD was defined as angina, MI, fatal CHD, and coronary revascularization. DbGap accession phs000287.v3.p1.

The Women's Health Initiative (WHI) [10]

A subset of the WHI cohort genotyped by Population Architecture using Genomic and Epidemiology (PAGE) study was also included for validation analyses. The WHI cohort is a long-term national health study with two main parts, namely the randomized clinical trial and observational study, that enrolled women ages 50-79 during 1993-1998 across 40 centers across the U.S. with follow up data available through 2018. Individuals with prevalent CHD at baseline were excluded and incident CHD was defined as MI including silent MI, as determined by ECG evidence and fatal CHD. Incident CHD events from all of the described NHLBI cohorts were clinically adjudicated. DbGap accession phs000200.v12.p3.

The electronic Medical Records and Genomics (eMERGE) Network [11,12]
The eMERGE network phases I-III cohort is comprised of participants from 12 contributing academic sites across the US with EHR-linked DNA biorepositories. Genotyping and phenotype data quality control of eMERGE dataset as well as the electronic phenotyping algorithm to ascertain incident CHD cases and controls have been previously described [13,14]. In brief, CHD cases were defined as the occurrence of either MI or a coronary revascularization event, such as percutaneous coronary intervention or coronary artery bypass grafting. Controls were defined as individuals without any diagnosis or procedure codes related to the CHD during EHR follow-up.

Subset of UKBB Cohort [15,16]
A subset of UKBB with only SAS participants were used to assess performance of the PRS determined via ADMIXTURE analysis. Briefly, in this study, more than 500,000 participants aged 40-70 were recruited between 2006 and 2010 from the general population through 22 assessment centers throughout the United Kingdom. UKBB individuals were genotyped using either the UK Biobank Axiom Array or the UK BiLEVE Axiom Array [17]. Incident CHD was ascertained based on CHD algorithm performance excluding prevalent cases at study enrollment, matching the eMERGE network.

dbGaP, eMERGE and UKBB
Data from dbGaP was downloaded via aspera/sraclient [18,19]. Each cohort was run through the Mayo GWAS QC pipeline to ensure only high-quality samples/SNPs were uploaded to the TOPMed imputation server [6,20,21]. Some cohorts had already undergone QC by their respective institutions, while others had all data provided regardless of quality. The pipeline was run in an iterative process that excluded samples/SNPs with poor genotype quality (< 95% Call Rates), samples with lower-than-expected heterozygosity (< 40%) on any chromosome, samples with F-statistic for sex check that differed from self-reported sex, and related subjects. Relationships were measured by King-Robust and relationships were identified as parent/offspring, full siblings, or duplicates [22,23]. One sample from each related pair was chosen at random to be removed. The program ADMIXTURE was used to estimate genetic ancestry admixture [24]. Genetic admixture was used to create homogenous groups (admixture probabilities > 75% for a single ancestry) to evaluate Hardy-Weinberg Equilibrium (HWE) [24]. Individuals with HIS ancestry were defined based on self-reported status, as it lacks a reference panel for use in ADMIXTURE. SNPs with HWE *p*-value $< 10^{-6}$ were removed.

After QC processing, samples were uploaded to the TOPMed imputation server where additional QC steps were completed, including removal of multi-allelic SNPs, indels, monomorphic SNPs, and SNPs with large allele frequency differences compared to Haplotype Reference Consortium (HRC)/1000G [6,20,21]. Table S1 displays detailed information on cohorts, including the total number of sample SNPs. Imputed SNPs with an imputation $R^2 > 0.3$ were retained.

Once imputation was completed, King-Robust was used to identify related subjects between different studies (i.e., ARIC and MESA), and one subject from each related pair was randomly removed [22,23]. 458,384 SNPs of the initial 542,218 SNPs from MVP $PRS_{P+T}$ passed QC and overlapped with 1000 Genomes reference panel, imputed dbGaP cohorts, eMERGE cohort, UKBB cohort, and the Global Diversity Array. The multi-ancestry $PRS_{CSx}$ had 626,955 SNPs of the initial 2,579,036 SNPs pass QC and overlap with the 1000 Genomes reference panel, imputed dbGaP, eMERGE, UKBB cohorts, and the Global Diversity Array. The imputed SNPs were used to calculate a PRS for each study participant using PLINK version 2.0 [23]. PRSs were computed as a weighted sum of dose of the risk allele, where weights were derived from the MVP summary statistics.

Biobank Japan (BBJ) [25,26] and Osaka Acute Coronary Insufficiency Study (OACIS)

In the East Asian validation analyses, the two Japanese cohorts [27] for validation included 1) BioBank Japan (BBJ) 2nd cohort: a hospital -based Japanese biobank data, excluding C4D meta-analysis and 2) OACIS cohort: a hospital-based registry where patients with acute myocardial infarction were enrolled. There were 14,827 samples (male 9841, female 4986; median age 69 [min 20, max 99]) of which there were 3,093 cases (279 from BBJ 2nd; 2,814 from OACIS) and 11,734 controls (from BBJ 2nd). Sample QC resulted in removal of several participants (1 high missingness; 2 PCA outliers, 101 sex mismatches, 5 missing clinical information).

Variants underwent QC to exclude SNPs with call rate > 0.99, HWE > 1e-06, and minor allele count ≥ 2. The remaining 503,561 variants were imputed to 20,322,240 variants with using the 1000 Genomes reference panel [4]. After QC, 458,385 variants were remaining that were used for the multi-ancestry $PRS_{P+T}$ validation and 1,120,251 variants available for the multi-ancestry $PRS_{CSx}$ validation.

The TAICHI Consortium [28]

The TAICHI cohort is a total of 7,924 Han Chinese individuals genotyped using the Illumina CardioMetaboChip v1.0 to identify genetic determinants of atherosclerosis and metabolic related traits [28]. Patients were enrolled in the study and defined as CHD positive if they meet one of the following criteria: history of myocardial infarction, history of coronary artery bypass graft or percutaneous coronary angioplasty and/or stenting, or at least 1 major vessel with stenosis ≥ 50% demonstrated by angiography [28]. Age of recruitment is the definition of age in this cohort. Sample QC includes removing individuals with high genotype missingness rate, extreme heterozygosity relatedness and cryptic relatedness with PI-Hat values > 0.125 and population outliers. Outliers were individuals with eigenvalues from PCs 1, 2, and 3 greater than 3 SDs from the mean [29].

SNPs are filtered for call rates < 98%, MAF < 5%, and HWE p-value < 0.05 [28]. A total of 466,450 variants were imputed to 292,323,483 variants using the NHLBI's TOPMed reference panel consisting of 97,256 deeply sequenced multi-ancestry human genomes [6,20]. 507,946 variants were used after QC for the $PRS_{P+T}$ validation and 1,288,975 variants were used for the $PRS_{CSx}$ validation.

#### Statistical Adjustments

dbGaP, eMERGE, and UKBB Cohorts

PRSs were centered and scaled by subtracting the mean and dividing by the SD [23]. Because the distribution of PRSs differed across different ancestries due to SNP allele frequency differences, we applied a continuous correction for population differences. PCA projection used 95,122 SNPs with 1000 Genomes for applying the Broad Institute method for genetic ancestry adjustment on raw PRS values computed for each participant. Following the approach by Khera et al. (2019) and suggestions by Christopher Kachulis, Broad Institute (personal communication), PRSs were corrected for genetic ancestry by a continuous correction based on projecting the study sample PRS onto the 1,000 Genomes reference sample [30]. This was accomplished by computing PRSs on the reference sample and using linear regression to regress the reference PRS on the top 4 principal components of the reference sample. The regression coefficients from this reference regression were used to predict the PRS in the study sample, by using the sample principal components. This predicted value was then subtracted from the sample PRS to adjust for each genetic ancestry. Because this approach only corrects for the mean of the distribution, it might not fully correct for genetic ancestry if the variance of the PRS differs across populations. To adjust for the variance, we created residuals from the linear regression in the reference sample, and then performed a second linear regression of the squared residuals on the components in the reference sample. This was then used to correct the variance in the study sample. To illustrate, the PRS adjusted score was computed as

$$PRS_{adjusted} = \frac{PRS_{sample} - (\alpha_0 + \sum \alpha_i PC_i)}{\sqrt{\beta_0 + \sum \beta_i PC_i}}$$

where α coefficients were estimated by regression of the PRS on the principal components in the reference samples, β coefficients were estimated by regression of the squared residuals on the principal components in the reference samples, and $PRS_{sample}$ and $PC_i$ are from the study samples (Figure S4).

BBJ, OACIS and TAICHI

Adjustments to the PRSs from these cohorts consisted of normalizing the scores for the mean and standard deviation of the genetic ancestry group. The multi-ancestry $PRS_{P+T}$ and $PRS_{CSx}$ validation in the BBJ and OACIS cohorts were adjusted together for age, sex and 10 PCs. The TAICHI cohort was separately validated and adjusted for age, sex, and 10 PCs. PRSs were then meta-analyzed using a fixed effect inverse-variance weighted model [31].

### Supplemental Table

Table S1. Studies obtained for PRS Validation

| Cohort study label | Data Access Information and study full name | Participants after QC (n) | | | | |
|---|---|---|---|---|---|---|
| | | EUR | AFR | HIS | EAS | SAS |
| Multi-population Meta-analysis GWAS (MVP, UKBB, CARDIoGRAMplusC4D, BBJ) | Tcheandjieu, C., Zhu, X., Hilliard, A.T. *et al.* Large-scale genome-wide association study of coronary artery disease in genetically diverse populations. *Nat Med* **28**, 1679–1692 (2022). https://doi.org/10.1038/s41591-022-01891-3 | 190,493 cases, 582,775 controls | 54,042 cases, 204,610 control | 24,234 cases, 89,706 controls | 29,319 cases, 183,134 controls | - |
| MVP Training | Million Veteran Program (MVP) | 24,041 cases, 32,318 controls | 3,980 cases, 13,007 control | 1,800 cases, 8,192 controls | 626 cases, 3,760 controls | |
| MVP validation | Million Veteran Program (MVP) | 6,158 cases, 61,580 controls | 1,552 cases, 15,520 control | 574 cases, 5,740 controls | - | - |
| The Atherosclerosis Risk in Communities Study (ARIC) | dbGaP (Study Accession# phs000280.v6.p1) under the project titled “Development and validation of polygenic risk score-based risk models for coronary heart disease in diverse racial/ethnic groups” | 1,696 cases, 6,629 controls | 465 cases, 1,805 control | - | - | - |
| Multi-Ethnic Study of Atherosclerosis (MESA) | dbGaP (Study Accession# phs000209.v13.p3) under the project titled “Development and validation of polygenic risk score-based risk models for coronary heart disease in diverse racial/ethnic groups” | 191 cases, 2,304 controls | 81 cases, 1,293 control | 72 cases, 1,079 controls | - | - |
| Cardiovascular Health Study (CHS) | dbGaP (Study Accession# phs000287.v7.p1) under the project titled “Development and validation of polygenic risk score-based risk models for coronary heart disease in diverse racial/ethnic groups” | 932 cases, 2,285 controls | 126 cases, 415 control | - | - | - |

| | | | | | | |
|---|---|---|---|---|---|---|
| Women's Health Initiative (WHI) | dbGaP (Study Accession# phs000200.v12.p1) under the project titled "Development and validation of polygenic risk score-based risk models for coronary heart disease in diverse racial/ethnic groups" | 32 cases, 313 controls | 396 cases, 5,403 control | 122 cases, 2,669 controls | - | - |
| eMERGE Phases I-III | version 3 imputed array data and accompanying phenotype data obtained directly from eMERGE Coordinating Center under the project NT406 titled "Improving Prediction for Coronary Heart Disease Risk Across Diverse Populations Using Transethnic Polygenic Risk Scores" | 2,119 cases, 42,427 controls | 291 cases, 7,484 control | 120 cases, 3,174 controls | - | - |
| Biobank Japan (BBJ) (2nd independent) cohort and OACIS | Hospital-based biobank excluding C4D meta-analysis, and Hospital-based registry of patients with AMI | - | - | - | 3,093 cases, 11,734 controls | - |
| UKBB | UK Biobank ADMIXTURE SAS individuals | - | - | - | - | 517 Cases 8,661 controls |
| TAICHI | TAICHI Consortium | - | - | - | 3,228 cases, 4,696 controls | - |

3

Table S2. Odds Ratios for incident CHD using PRS-CSx derived weights from EUR and multi-ancestry based GWAS effect size estimates in diverse ancestry cohorts.

| **Ancestry** | **Age [a] (mean±SD[b])** | **Cases/Controls** | **PRS-CSx GWAS Summary Statistics** | **AUC[c]** | **OR[d] (95% CI[e]) per 1 SD** | ***P*-value** | **OR (95% CI) Top 5% vs Rest** | ***P*-value** |
|---|---|---|---|---|---|---|---|---|
| EUR | 52.4 ± 15.5 | 4,970/47,732 | EUR | 0.773 | 1.55 (1.50-1.60) | 4.51E-164 | 2.40 (2.15-2.69) | 2.34E-53 |
| | | | Multi | 0.774 | 1.65 (1.59-1.71) | 5.19E-171 | 2.48 (2.23-2.77) | 5.88E-61 |
| AFR | 53.4 ± 14.7 | 1,359/15,649 | EUR | 0.734 | 1.25 (1.17-1.33) | 8.98E-12 | 1.71 (1.37-2.13) | 1.78E-06 |
| | | | Multi | 0.736 | 1.20 (1.15-1.26) | 7.46E-14 | 1.74 (1.41-2.15) | 3.03E-07 |
| HIS | 54.8 ± 14.3 | 314/5,824 | EUR | 0.708 | 1.52 (1.36-1.71) | 1.75E-12 | 2.04 (1.36-3.05) | 5.24E-04 |
| | | | Multi | 0.706 | 1.51 (1.35-1.69) | 7.48E-13 | 2.57 (1.77-3.73) | 7.98E-07 |
| EAS | 61.0 ± 14.8 | 3,228/4,696 | EUR | 0.756 | 1.51 (1.44-1.59) | 5.80E-55 | 1.93 (1.58-2.36) | 1.49E-10 |
| | | | Multi | 0.762 | 1.59 (1.54-1.64) | 2.41E-160 | 2.34 (2.06-2.66) | 1.78E-28 |
| SAS | 53.2 ± 8.4 | 517/8,661 | EUR | 0.803 | 2.47 (2.23-2.73) | 3.17E-67 | 5.36 (4.15-6.92) | 7.91E-38 |
| | | | Multi | 0.803 | 2.67 (2.39-3.01) | 1.48E-63 | 4.92 (3.81-6.35) | 3.90E-34 |

[a] Age- Age at enrollment
[b] SD- Standard Deviation
[c] AUC-Area under the Curve
[d] OR- Odds Ratio
[e] CI- Confidence Interval

#### Supplemental Figures

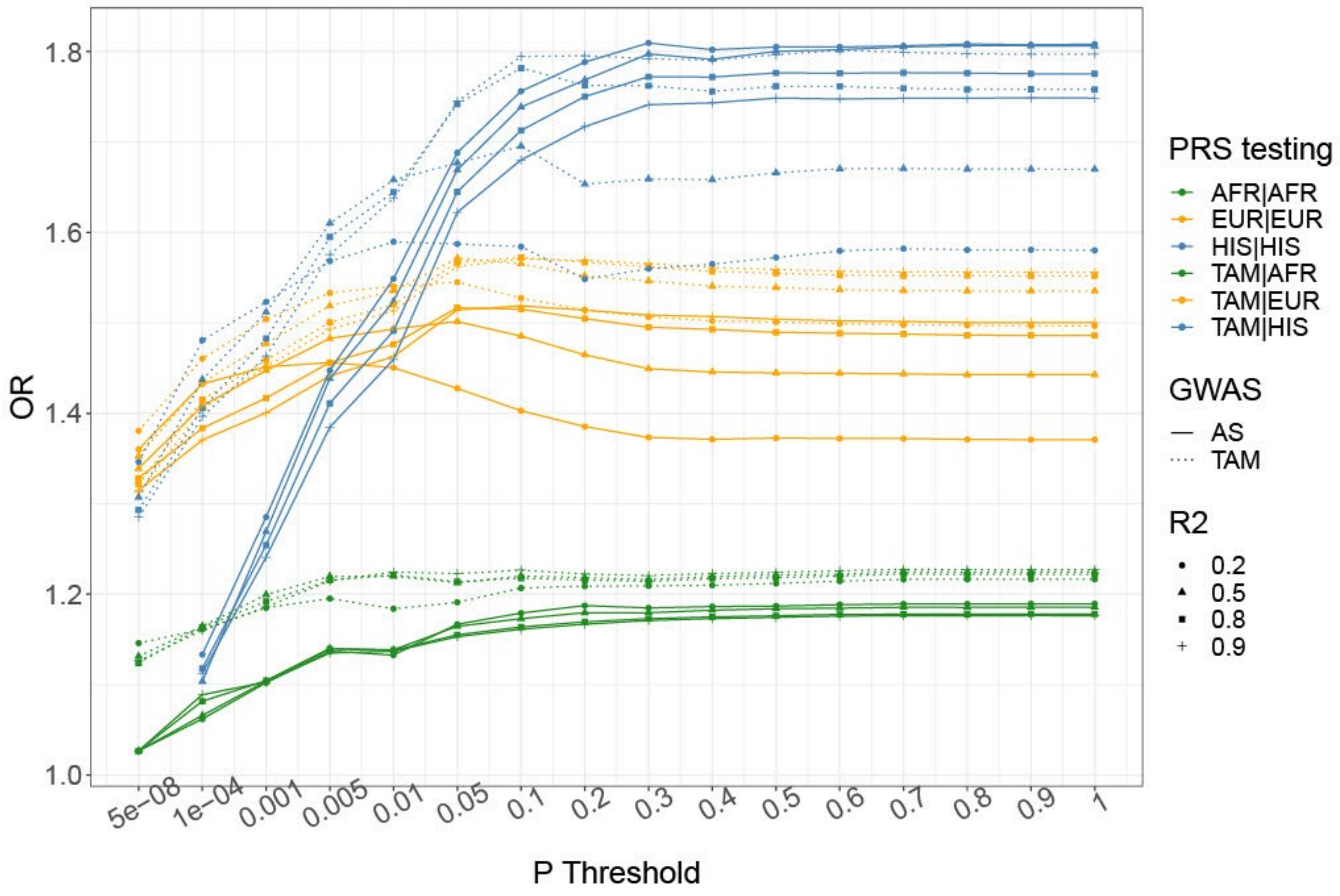


Figure S1. Plot describing the PRS testing using P+T method across European (EUR), African (AFR), and Hispanic (HIS) genetic ancestry populations. Each color represents the population in which the GWAS was derived (except for the meta-analysis) and PRS tested. The solid line represents the PRS testing using ancestry specific (AS) GWAS, the dash line represents the PRS testing using the meta-analysis or trans-ancestry (TAM) GWAS.

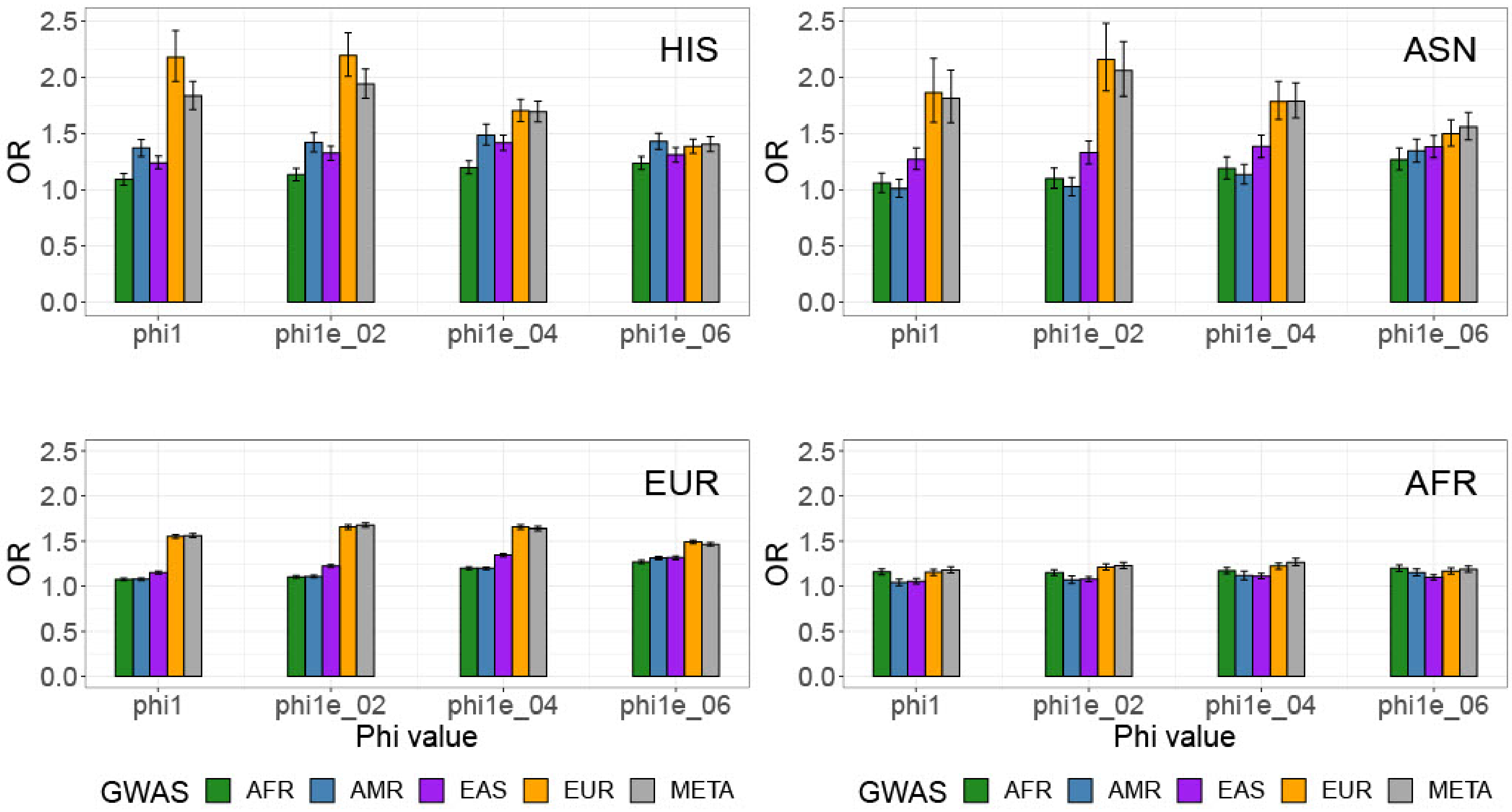


Figure S2. Plot describing the PRS testing using continue shrinkage method across European (EUR), African (AFR), and Hispanic (HIS) genetic ancestry populations. Each color represents the population in which the GWAS was derived.

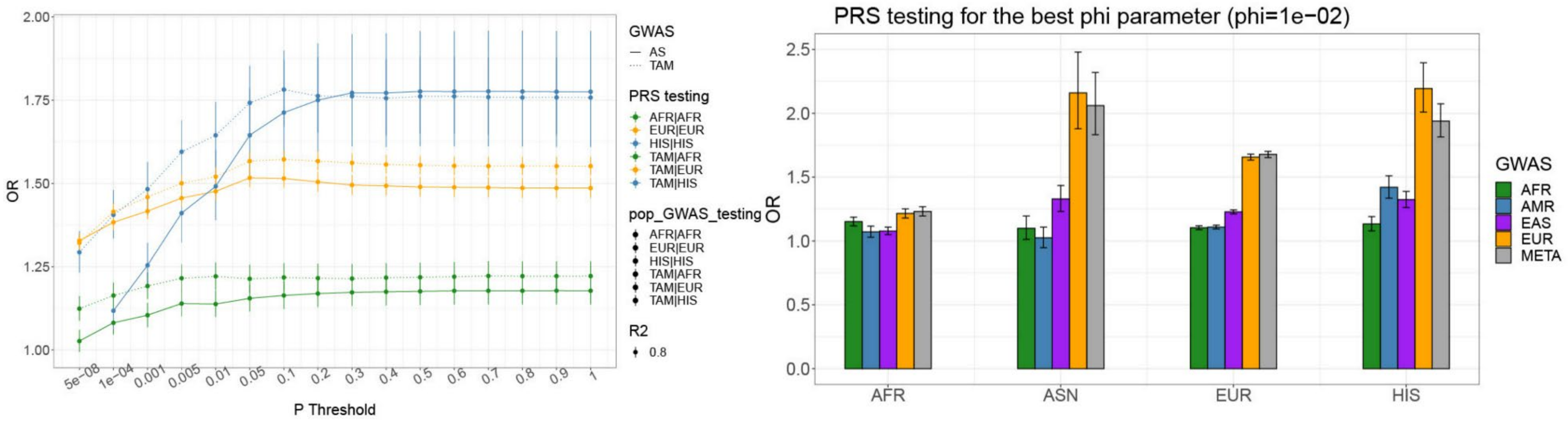


A.) B.)

Figure S3. (A) Plot performance PRS testing for the best R2 (R2 ≤ 0.8) and best windows (250 kb) for the pruning and thresholding methods. Each color represents the population in which the GWAS was derived (for the solid line) and PRS tested. The solid line represents the PRS testing using ancestry specific GWAS summary statistic, the dash line represents the PRS testing using the meta-analysis summary statistic. (B) Plot PRS performance comparing the prediction of various PRS for the best phi parameter when using continue shrinkage methods (PRS-CSx).

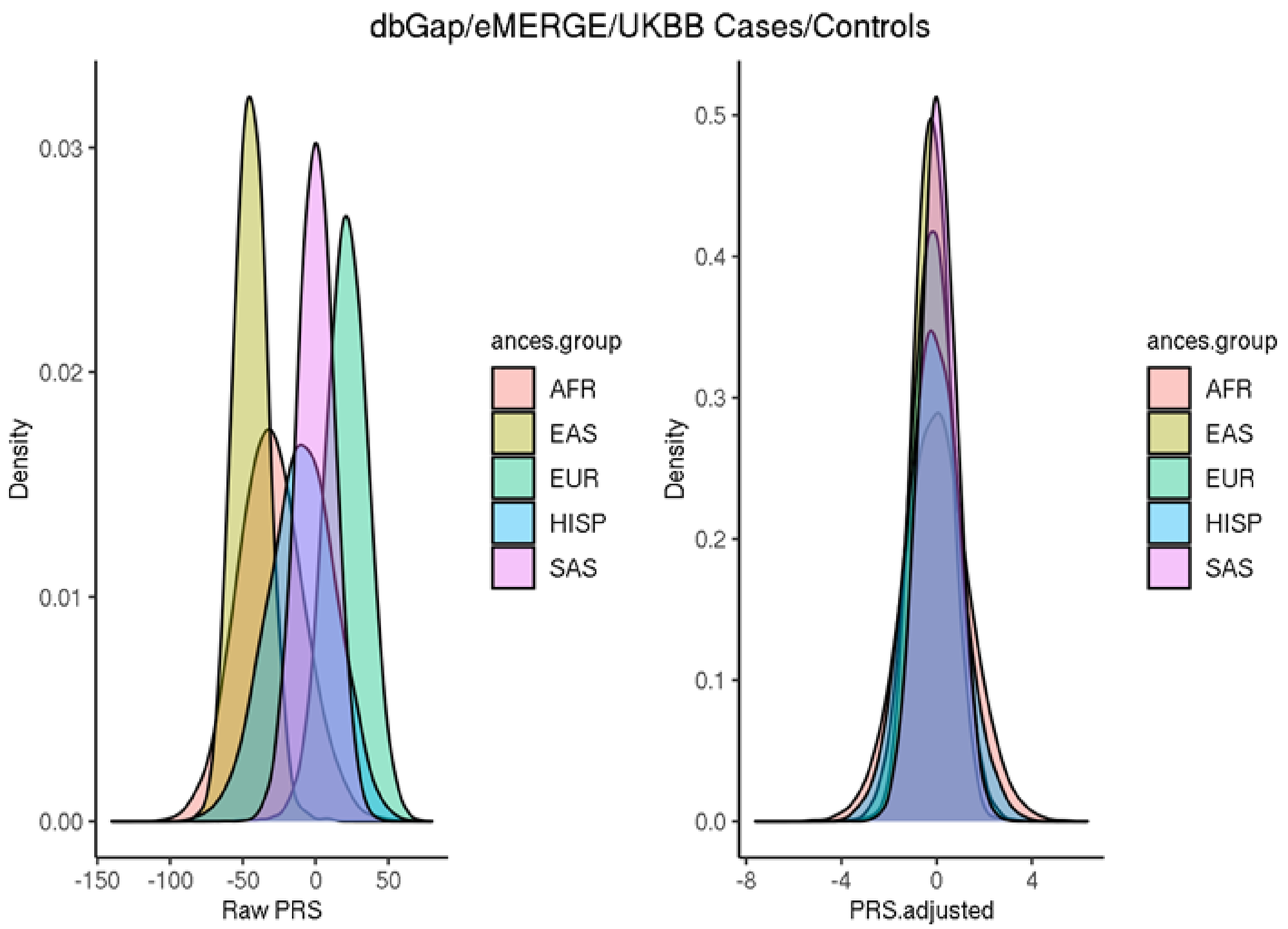


Figure S4. Distributions of raw and adjusted PRS according to genetic ancestry groups.

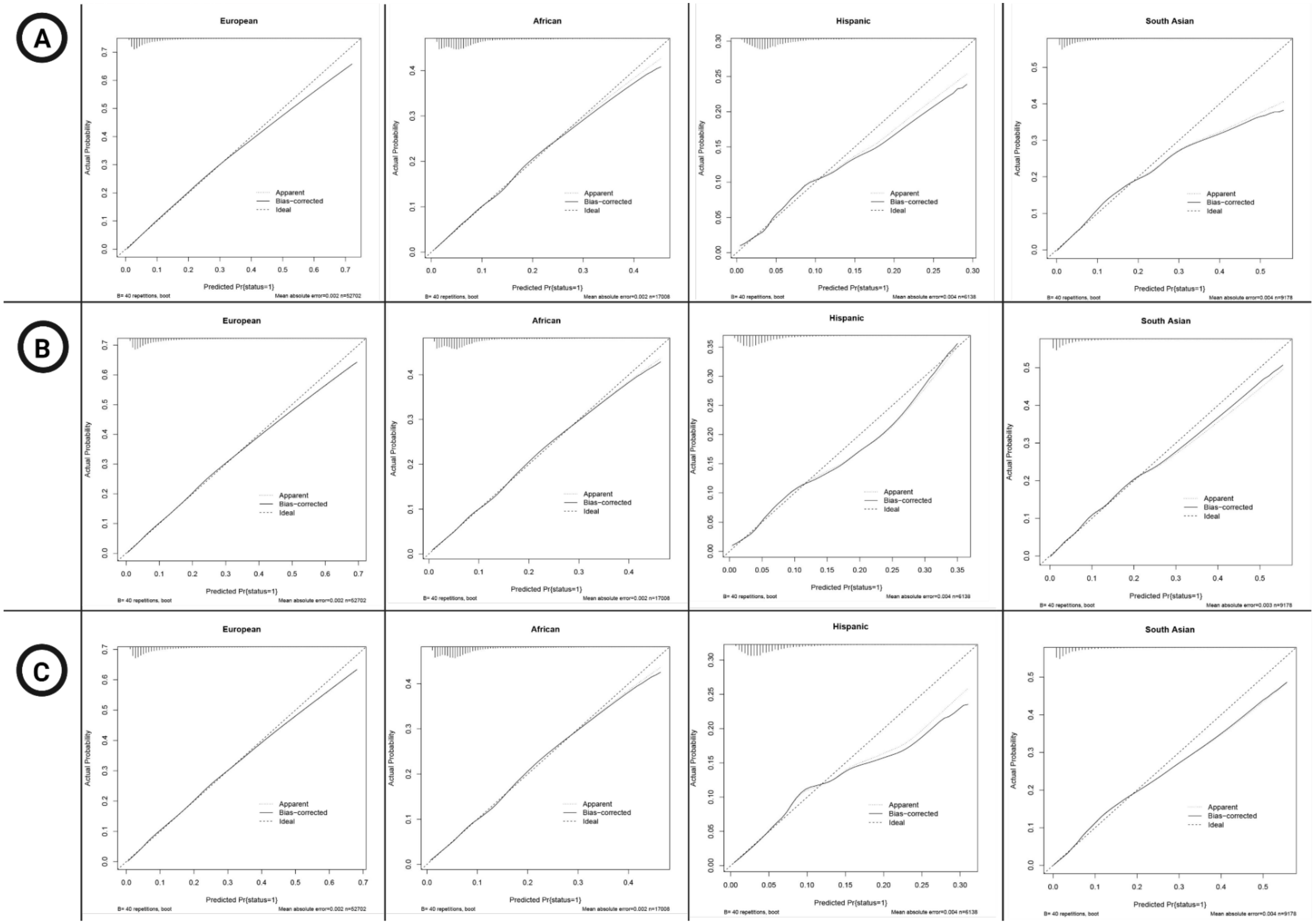


Figure S4. Calibration plots of the best-performing multi-ancestry (A) multi-ancestry $PRS_{P+T}$ and (B) multi-ancestry $PRS_{CSx}$ and (C) EUR-trained $PRS_{CSx}$ for European (EUR), African (AFR), Hispanic (HIS), and South Asian (SAS) genetic ancestry populations based on external validations from dbGaP and eMERGE cohorts.
